# Comparison of videolaryngoscope & direct laryngoscope for intubation in critically ill patients: A Bayesian meta-analysis

**DOI:** 10.1101/2020.04.21.20074153

**Authors:** Souvik Maitra, Anirban Som, Sulagna Bhattacharjee

## Abstract

**Purpose:** To identify the benefit of video laryngoscope (VL) over direct laryngoscope (DL) for intubation in the intensive care unit (ICU)

**Material & Methods:** Randomized controlled trials (RCTs) comparing VL with DL for intubation in ICU by was conducted in conventional frequentist methodology and also incorporated of the previous evidences from observational studies in Bayesian methodology.

**Results:** Data of 1464 patients from six RCTs have been included in this meta-analysis. In conventional meta-analysis of RCTs, first attempt intubation success rate was similar between VL and DL group [p=0.39]. Rate of esophageal intubation was significantly less with VL [p=0.03] and glottic visualization was significantly improved with VL in comparison to DL [p=0.009]. Time to intubation was similar in both the group [p=0.48]. When evidences from a meta-analysis of observational studies incorporated in Bayesian model, first attempt intubation success is significantly higher with VL [posterior median log OR (95% credible interval) 0.50 (0.06, 1.00)].

**Conclusion:** Evidences from both observational studies and RCTs synthesized in Bayesian methodology suggest that use of VL for endotracheal intubation in critically patients may be associated with higher first intubation success when compared to DL.

---

Emergency endotracheal intubation are often required in critically ill patients for providing ventilatory support and airway protection. More than half of the airway related adverse events in the intensive care unit is associated with death or permanent neurological injury^1^. Airway events in the ICU or emergency department happened more in the odd hours and managed by physicians with less experience in anesthesiology leading severe consequences. Laryngoscopy attempt of more than two during emergency intubation was associated with higher incidence of hypoxemia, regurgitation of gastric content, aspiration, bradycardia and cardiac arrest^2^.

Video laryngoscopy improves glottic visualization and associated with less failed intubation particularly in patients with difficult airway^3,4^. Clinical usefulness of video laryngoscope in emergency scenarios such in the ICU or emergency department is less clear. Huang et al, in a meta-analysis of randomized controlled trials reported that video laryngoscopy does not offer any advantages such as first attempt intubation success rate, time to intubation, complications and mortality for emergency intubation in critically ill patients^5^. Utility of VL in the ED is controversial; though observational studies reported benefit, a meta-analysis of randomized controlled trial reported no benefit of video laryngoscopy in terms of first attempt success rate or overall intubation success rate during intubation in the ED^6^. However, a lower esophageal intubation rate was reported in that meta-analysis.

A recent meta-analysis by Arulkumaran et al in 2018^7^, reported that VL is associated with higher first attempt intubation rate in the ICU, but not in the ED or in prehospital setting. In that meta-analysis, data from both randomized controlled trials (RCTs) and observational studies polled in a single analysis, which might have incurred significant bias. In this present meta-analysis, evidences from observational studies have been incorporated in the evidences from randomized controlled trials as per Bayesian methodology.

## Methods

We followed the recommendations of Preferred Reporting Items for Systematic Reviews and Meta-Analyses (PRISMA) statement for conducting and reporting the results of this meta-analysis^8^. Protocol of this meta-analysis was registered at the International Prospective Register of Systematic Review (PROSPERO, CRD42019131618).

### Eligibility criteria

Randomized controlled trials comparing any form of video laryngoscopy such as C-MAC^®^ or glidescope^®^ or MacGrath^®^ videolaryngoscope etc. with direct laryngoscopy in adult patients requiring endotracheal intubation in the intensive care unit has been included in this meta-analysis. We also searched for the both prospective and retrospective observational studies reported first attempt intubation success with VL compared to DL to formulate a ‘prior information’ for incorporation in this meta-analysis as per Bayesian methodology.

### Information sources & search strategy

PubMed (1946 to 15^th^ May 2019), EMBASE and The Cochrane Central Register of Controlled Trials (CENTRAL) were searched for potentially eligible trials on 15^th^ May 2019. References of the previously published meta-analyses and randomized controlled trial were also hand searched for eligible trials. Following keywords were used to search database: ‘video laryngoscopy’, ‘video laryngoscope’, ‘direct laryngoscopy’, ‘direct laryngoscope’, ‘C-MAC’, ‘glidescope’, ‘McGrath video laryngoscope’, ‘ICU, ‘intensive care unit’, ‘critical care unit’ ‘critically ill’. A similar search strategy was used by Bhattacharjee et al previously^6^.

### Study selection

Two authors (SM and SB) independently searched title and abstract of the potentially eligible articles to be included in this meta-analysis. Finally, full texts of the possible articles were retrieved and assessed for eligibility. Any disputes between the two authors were solved by consultation with the third author (AS).

### Data collection process

Two authors (SM & SB) independently retrieved required data from the eligible RCTs and observational studies. All data were initially tabulated in a Microsoft Excel(tm) (Microsoft Corp., Redmond, WA) data sheet. Another author crosschecked these data before analysis (AS).

### Data items

Following data were retrieved from the full text for all studies: first author, year of publication, sample size, characteristics of included patients, first attempt intubation rate, overall intubation rat, time to intubate, rate of esophageal intubation and any other reported complications.

### Risk of bias in individual studies

Two authors (AS & SB) independently assessed the methodological quality of the included studies. Following methodological questions were searched from the studies as per the Cochrane methodology: method of randomization, allocation concealment, blinding of the participants and personnel, blinding of outcome assessment, incomplete data reporting, selective reporting and any other bias. For each area of bias, we designated the trials as low risk of bias, unclear risk of bias or high risk of bias. Risk of bias at individual study level will be graphically presented in the review.

### Summary measures and synthesis of results

Primary outcome of this meta-analysis was ‘first attempt intubation rate’ in the included patients. Secondary outcomes are overall intubation success rate, oesophageal intubation rate, time to intubation and any other reported complications related to the device use.

For continuous variables, mean and standard deviation (SD) values were extracted for both groups, a mean difference was computed at the study level, and a weighted mean difference was computed in order to pool the results across all studies. If the values were reported as median and an inter-quartile range or total range of values, the mean value was estimated using a previously described methodology^9^.

Odds ratio (OR) with 95% confidence interval was calculated for each trial and the pooled OR was estimated using the inverse variance method. The Q-test was used to analyse heterogeneity of trials and I^2^ statistics was reported. Considering possible clinical heterogeneity due to study design and patients’ population, we used a random effect model for all pooled analysis and STATA version 13.0 for Mac OS X was used for all frequentist analysis *(STATA SE 13*.*0, Stata Corp, College Station, TX, USA)*. Publication bias was assessed by visual inspection of funnel plot. For Bayesian random effect meta-analysis, we used ‘*bayesmeta’* package of **R** (***R*** *Core Team (2013)*. ***R***: *A language and environment for statistical computing*. ***R*** *Foundation for Statistical Computing, Vienna, Austria*.) within normal-normal hierarchical model. We planned to use improper, non-informative and informative µ (effect size). We also planned to use both proper (half normal distribution) and improper (Jeffrey’s prior) for heterogeneity prior (*τ)*. Prior effect size was determined from a meta-analysis of the observational studies and log of the effect size and the standard error of the log effect size was noted. As the pooled data derived from observational studies are possibly less robust, a weakly informative prior model will be also be used. Posterior marginal effect size will be reported as median of log OR with 95% credible interval and posterior predictive p value derived by Markov Chain Monte Carlo method.

## Results

Initial database searching and other sources revealed 4326 articles and after duplicate removal 2452 unique articles were identified. Amongst these articles, 85 articles were assessed from abstract and full text and six randomized controlled trials were included in this meta-analysis^10-15^. A PRISMA flow diagram showing selection of trials have been depicted in figure 1. Risk of biases as per Cochrane methodology has been depicted in figure 2. We have also found six observational studies, where first attempt intubation success was reported^16-21^.

**Figure 1:**
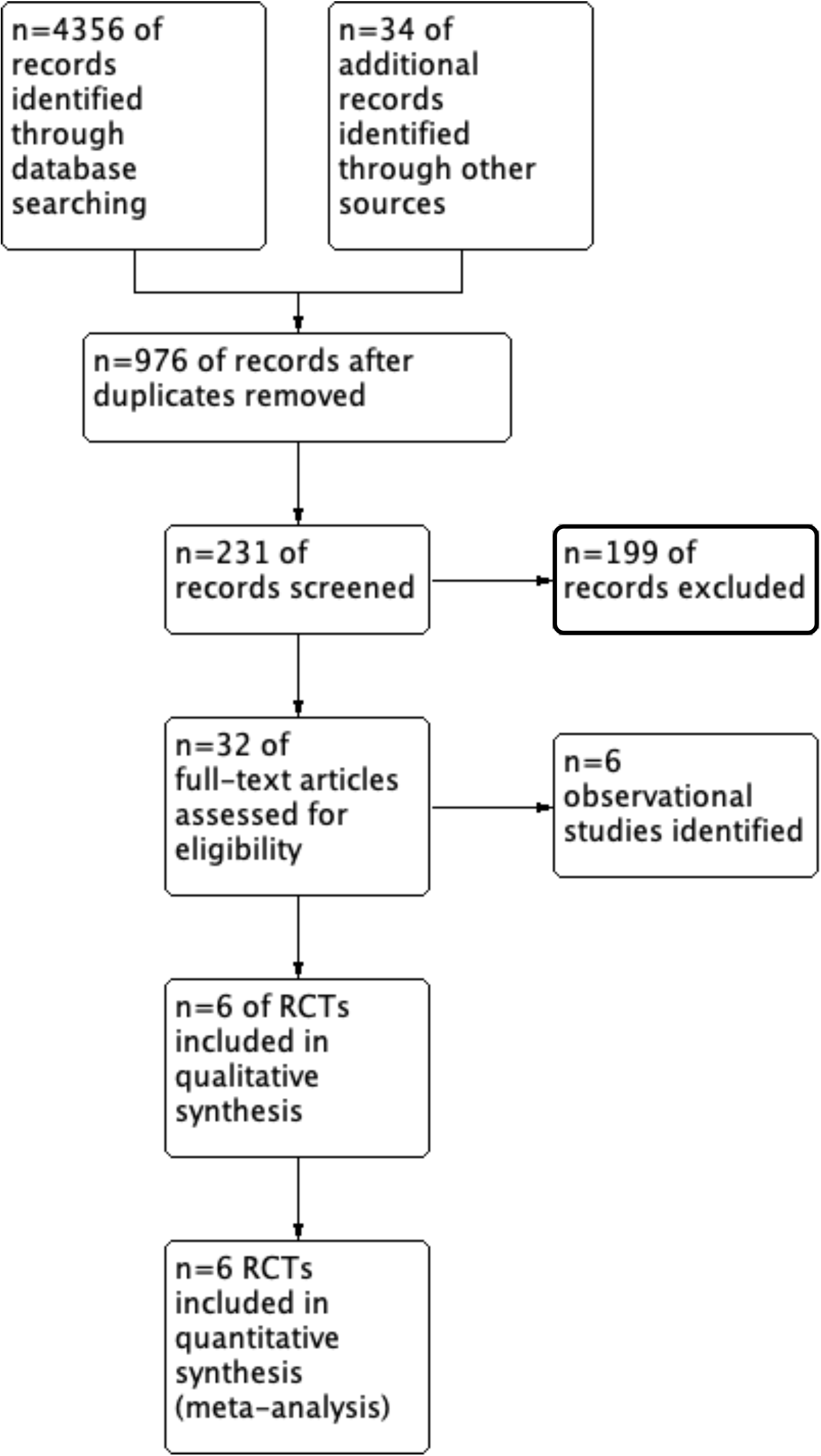
PRISMA flow diagram showing steps in literature search and study selection

**Figure 2:**
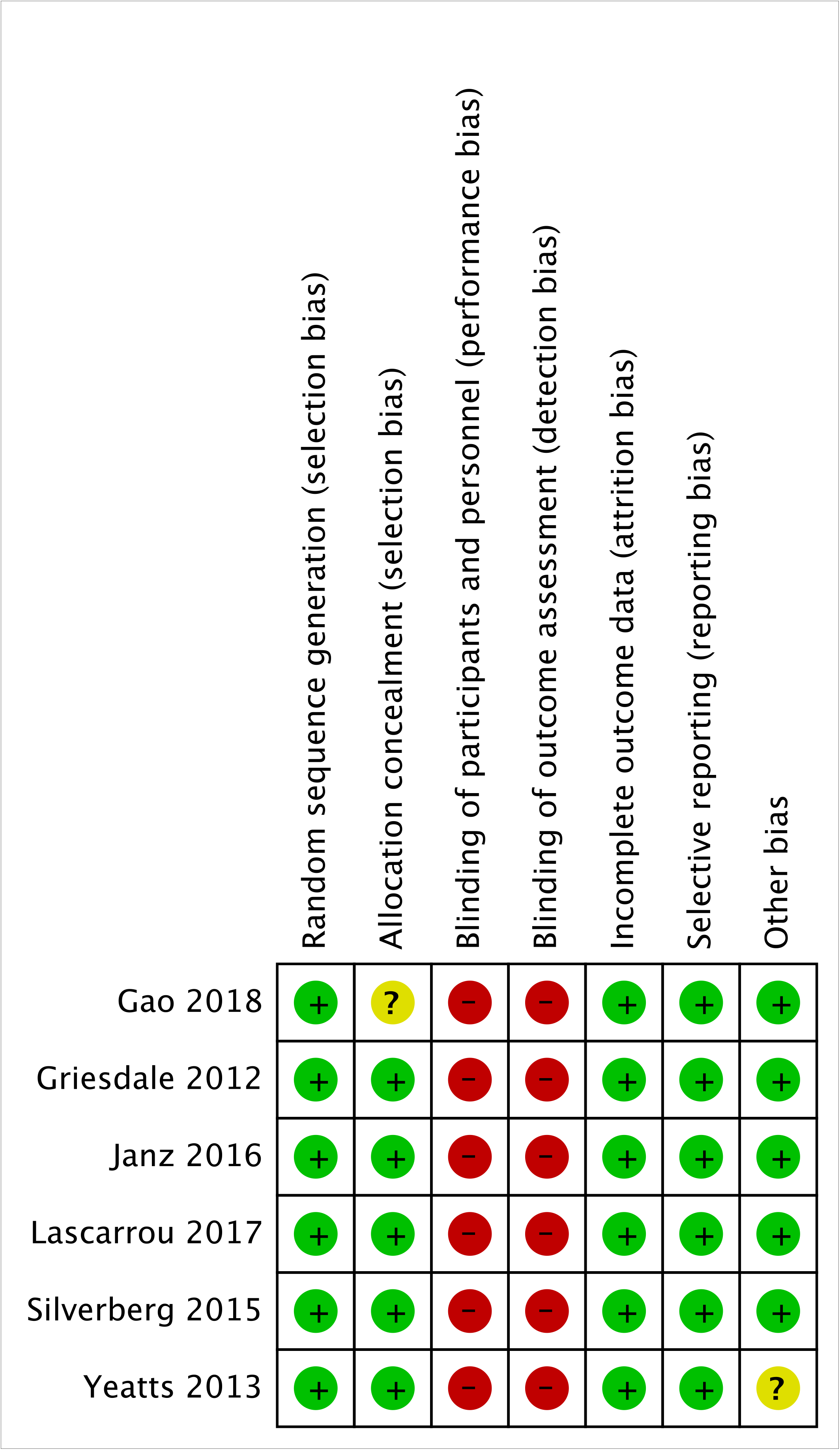
Risk of bias summary: review authors’ judgements about each risk of bias item for each included study.

### Conventional Meta-Analysis of the RCTs

All six RCTs reported first attempt intubation success rate and it was similar between VL and DL group [OR (95% CI) 1.19 (0.80, 1.76); inverse variance random effect model, p=0.39, I^2^=57.8%]. Four RCTs reported rate of esophageal intubation and it was found to be significantly less with VL [OR (95% CI) 0.38 (0.16-0.92); inverse variance random effect model, p=0.03, I^2^=0%]. However, glottic visualization was significantly improved with VL in comparison to DL [OR (95% CI) 2.97 (1.32, 6.70); inverse variance random effect model, p=0.009, I^2^=68.1%]. Three studies reported ‘all complications’ and it was found to be similar in two laryngoscopy methods [OR (95% CI) 0.79 (0.50, 1.26); inverse variance random effect model, p=0.32, I^2^=39.8%]. Time to intubation was similar in both the group [MD (95% CI) 7 (−12, 26) s, p=0.48, I^2^=95.5%]. A summary of forest plots of different outcomes has been provided in figure 3. Sensitivity analysis by excluding the RCT by Yeatts et al., as it was conducted in trauma resuscitation unit, also revealed similar first attempt intubation success rate.

**Figure 3:**
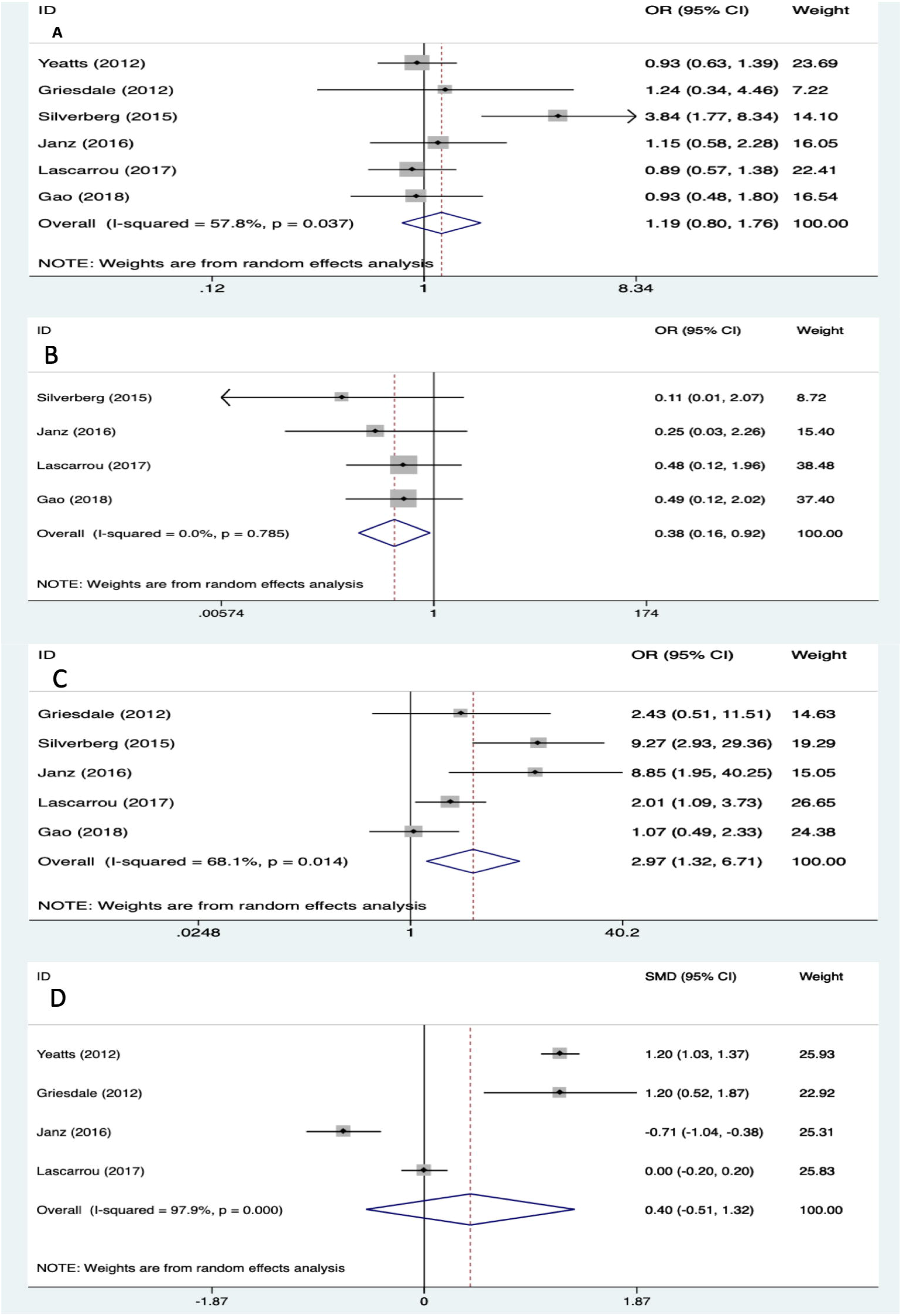
Forest plot showing pooled effect of video laryngoscopy on (A) odds ratio of first attempt intubation success, (B) odds ratio of esophageal intubation, (C) odds ratio of improved glottic view, (D) standard mean difference of time to intubation.

### Evidences from observational studies

A random effect inverse variance meta-analysis of 5 observational studies found that VL is associated with significantly higher first attempt success rate [log OR (95% CI) 0.90 (0.66-1.13)], similar incidence of complications [log OR (95% CI) −0.49 (−1.14, 0.17)] and rate of esophageal intubation [log OR (95% CI) −1.68 (−3.78, 0.41)].

### Bayesian random effect meta-analysis

With improper *µ and proper τ* prior (half normal distribution), posterior median log OR for first attempt intubation success was 0.16 with a 95% credible interval of *-0*.*35 to 0*.*75*. Log of effect size obtained from the meta-analysis of observational studies was 0.9 with a SD of 0.1. As evidences obtained from observational studies are prone to several biases, we conservatively assumed that true SD may be as high as 0.3. So, with an informative *µ* prior and *proper τ* prior (half normal distribution), posterior median log OR for first attempt intubation success was 0.50 with a 95% credible interval of *0*.*06 to 1*.*00*. However, with a non-informative *µ and proper τ* prior (half normal distribution), posterior median of *0*.*16* with a 95% credible interval of *-0*.*35 to 0*.*75 was obtained*. With an improper *µ* and improper *τ* (Jeffrey’s prior), posterior median log OR is 0.17 and a 95% credible interval of *-0*.*36 to 0*.*77*. With an informative *µ* prior and improper *τ* prior (Jeffrey’s prior), posterior median log OR for first attempt intubation success was 0.50 and a credible interval of 0.07 to 1.01. However, no benefit of VL was seen when a non-informative *µ* prior was used. Summary results from different priors have been provided in table 2. Figure 4 depicts forest plots and figure 5 depicts joint posterior density of effect size and heterogeneity parameter of the all six Bayesian model used here.

**Table 1:**
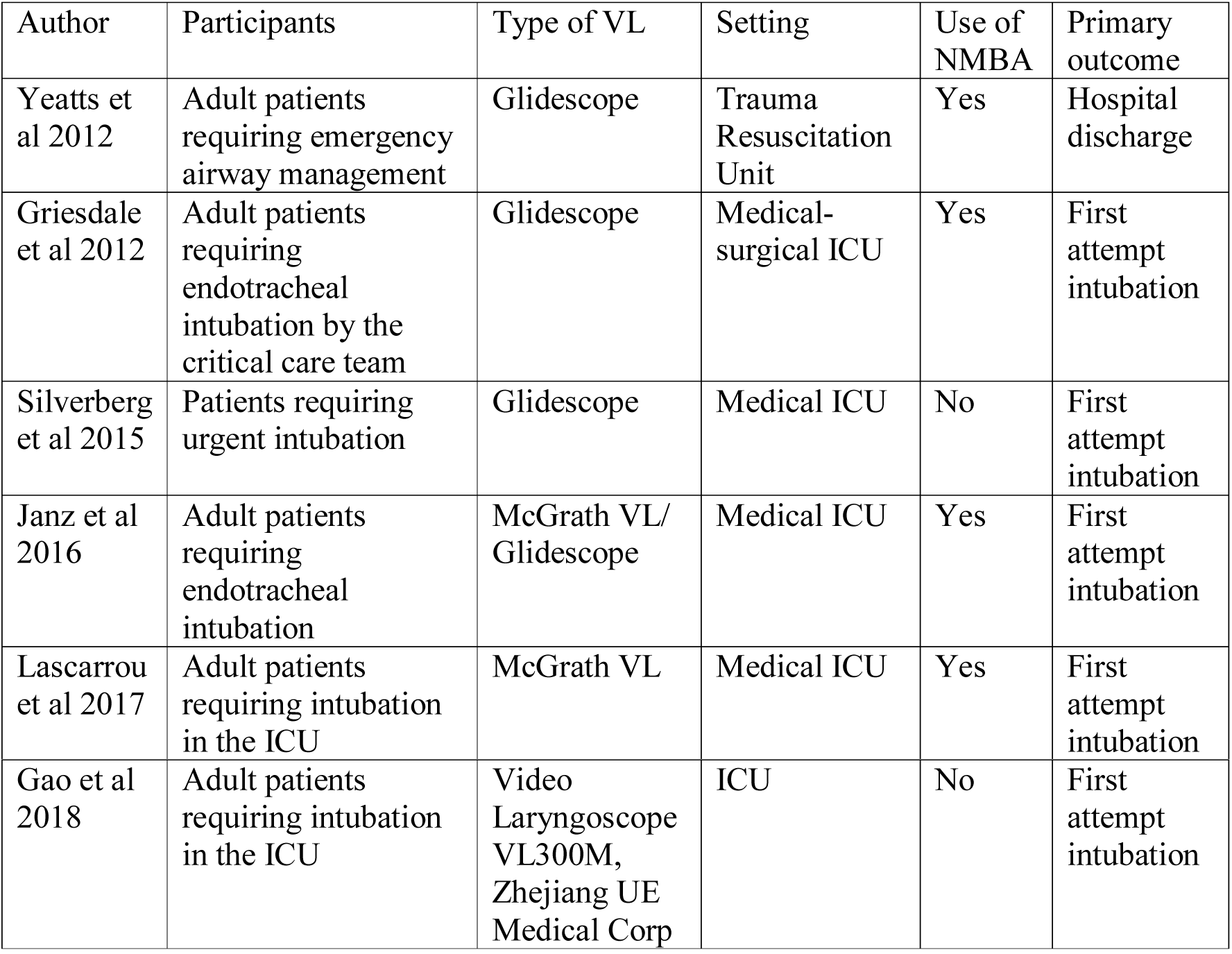
Characteristics of the included studies

**Table 2:**
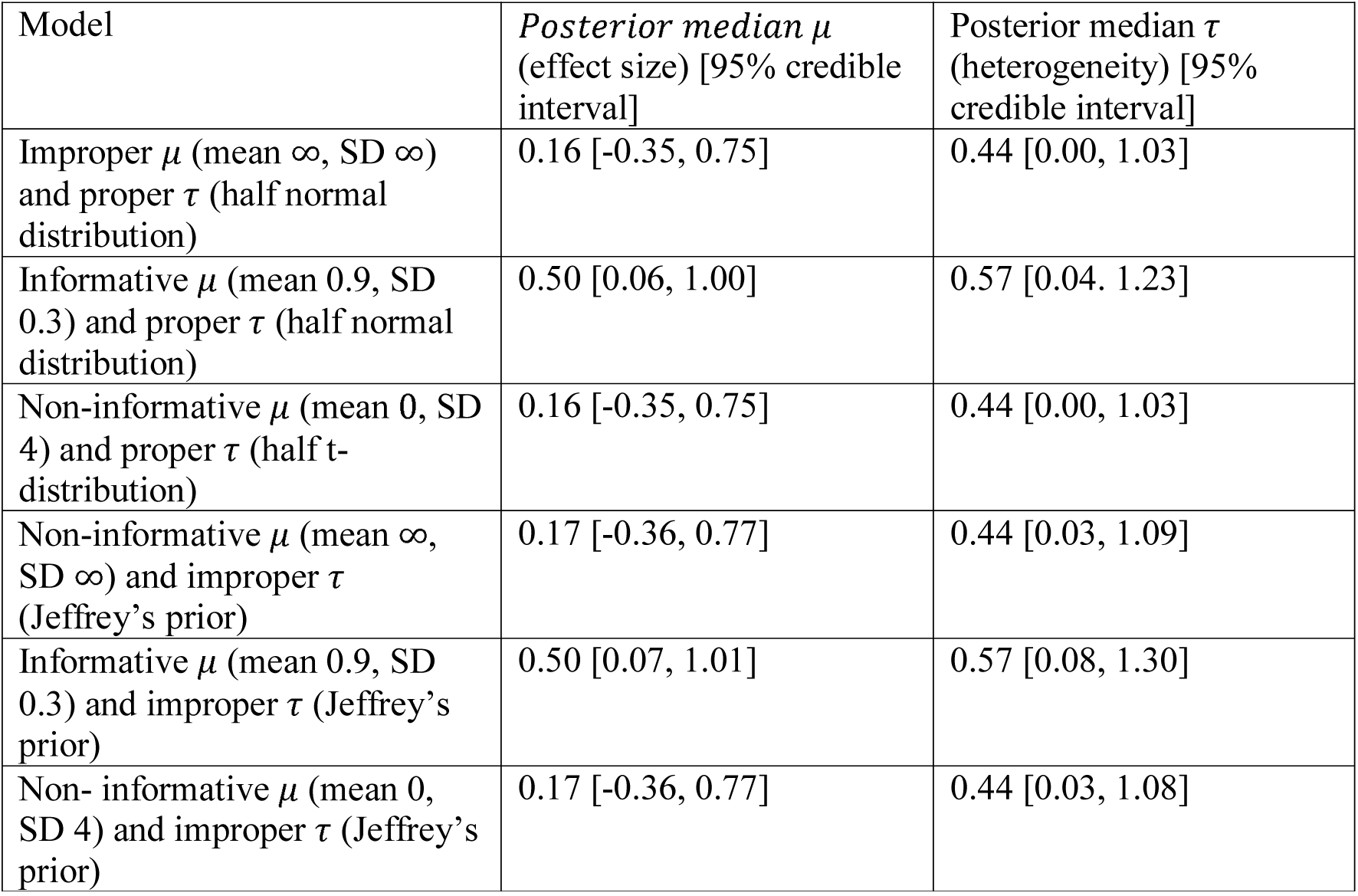
Posterior marginal values of median effect size and heterogeneity for first intubation success rate with different priors

**Figure 4:**
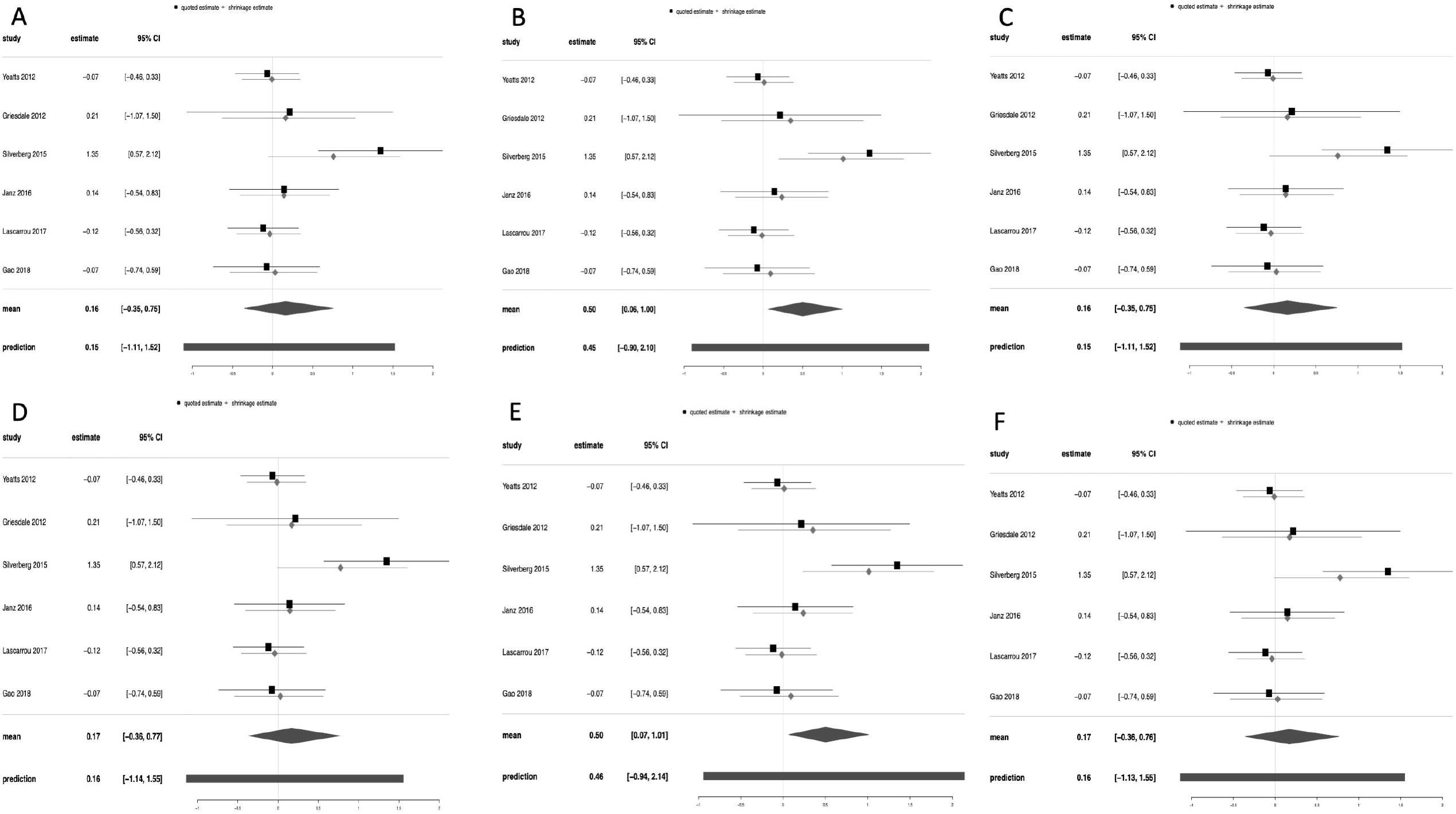
Forest plot showing posterior median with 95% credible interval of log odds ratio of first attempt intubation success from the following models: (A) Improper *µ* (mean ∞, SD ∞) and proper *τ* (half normal distribution); (B) informative *µ* (mean 0.9, SD 0.3) and proper *τ* (half normal distribution); (C) non-informative *µ* (mean 0, SD 4) and proper *τ* (half t- distribution); (D) non-informative *µ* (mean ∞, SD ∞) and improper *τ* (Jeffrey’s prior); (E) informative *µ* (mean 0.9, SD 0.3) and improper *τ* (Jeffrey’s prior); (F) non-informative *µ* (mean 0, SD 4) and improper *τ* (Jeffrey’s prior).

**Figure 5:**
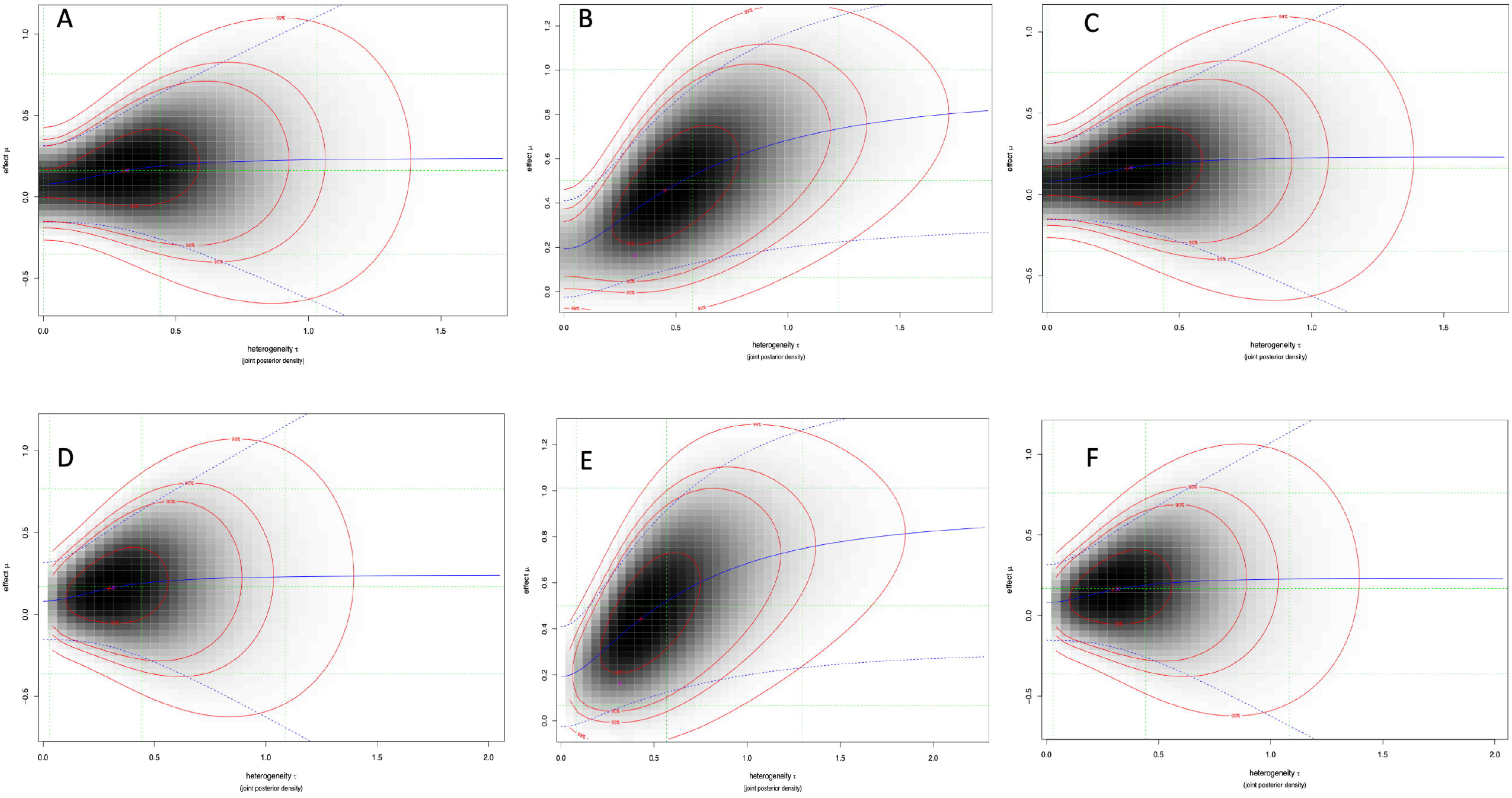
Joint posterior distribution of heterogeneity parameter (*τ*) and effect size (*µ*) for log OR of first attempt intubation success (A) Improper *µ* (mean ∞, SD ∞) and proper *τ* (half normal distribution); (B) informative *µ* (mean 0.9, SD 0.3) and proper *τ* (half normal distribution); (C) non-informative *µ* (mean 0, SD 4) and proper *τ* (half t- distribution); (D) non-informative *µ* (mean ∞, SD ∞) and improper *τ* (Jeffrey’s prior); (E) informative *µ* (mean 0.9, SD 0.3) and improper *τ* (Jeffrey’s prior); (F) non-informative *µ* (mean 0, SD 4) and improper *τ* (Jeffrey’s prior).

## Discussion

Principal finding of this meta-analysis is use of VL during endotracheal intubation in critically ill patients is associated with significantly higher first attempt intubation success in comparison to DL when evidence from prior observational studies are incorporated in the evidences from the randomized controlled trials. Glottic visualization during laryngoscopy is significantly improved and rate of esophageal intubation is also significantly decreased with VL.

Visualization of glottis during direct laryngoscopy requires the oral, pharyngeal, and laryngeal axes to be brought into a straight line, whereas glottic visualization using a videolaryngoscope does not^22^. American Society of Anaesthesiologists’ suggests that a good laryngeal exposure is one of the most important factors for successful intubation^23^. So, VL may be associated with higher first attempt intubation success compared to DL. A previous Cochrane review also reported that glottic visualization is improved with VL in adult patients^4^.

Randomized controlled trials are not unanimous in reporting superiority of VL over DL for endotracheal intubation in critically ill patients. A previous meta-analysis of 5 RCTs that included around 1300 patients reported that VL offers no advantage over DL in terms of first attempt intubation success rate^5^. However, another meta-analysis of both RCTs and observational studies in 2018 reported that VL offers higher first intubation success rate for emergency intubation in critically ill patients^7^. Authors of that meta analysis pooled the effect estimate both from RCTs and observational studies in same analysis, which had probably incurred significant amount of bias. Our conventional meta-analysis of RCTs reported that first intubation success rate is similar between VL and DL; however, incorporating prior evidences from observational studies even in a conservative manner indicated that VL is associated with higher first intubation success rate. It is beyond doubt that VL improves glottic view, but whether improved glottic visualization translates into a higher intubation success is debatable2^4^, possibly because of the view obtained in video-laryngoscope is indirect from a camera located at the tip of the laryngoscope blade.

### Strength & limitations

Most important strength of this meta-analysis is the use of Bayesian methodology for incorporating prior evidences from observational studies which was suggested by Sutton & Abrams^25^. Bayesian approaches to meta-analysis also allows full uncertainty for prediction. Most important limitation is the subjective interpretation of the prior effect size; though we have accepted a conservative standard deviation of the prior effect size, it still may be biased. Another important limitation of our meta-analysis is that success of endotracheal intubation may be variable in different video laryngoscope device and also according to the expertise of the physicians. As the number of the included trials are small, a meta-regression analysis was not possible here.

## Conclusion

Evidences from both observational studies and RCTs synthesized in Bayesian methodology suggest that use of VL for endotracheal intubation in critically patients may be associated with higher first intubation success when compared to DL.

## Data Availability

Data will be available from the authors

